# A quantitative evaluation of aerosol generation during manual facemask ventilation

**DOI:** 10.1101/2021.08.23.21262441

**Authors:** A. J. Shrimpton, J. M. Brown, F. K. A. Gregson, T. M. Cook, D.A. Scott, F. McGain, R. S. Humphries, R. S. Dhillon, B. R. Bzdek, F. Hamilton, J. P. Reid, A. E. Pickering, on behalf of the AERATOR study group

## Abstract

Manual facemask ventilation, a core component of elective and emergency airway management, is classified as an aerosol generating procedure. This designation is based on a single epidemiological study suggesting an association between facemask ventilation and transmission from the SARS 2003 outbreak. There is no direct evidence to indicate whether facemask ventilation is a high-risk procedure for aerosol generation. We conducted aerosol monitoring during routine facemask ventilation, and facemask ventilation with an intentionally generated leak, in anaesthetised patients with neuromuscular blockade. Recordings were made in ultraclean theatres and compared against the aerosol generated by the patient’s own tidal breathing and coughs. Respiratory aerosol from tidal breathing was reliably detected above the very low background particle concentrations (191 (77-486 [3.8-1313]) versus 2.1 (0.7-4.6 [0-12.9] particles.l^-1^ median(IQR)[range], n=11, p=0.002). The average aerosol concentration detected during facemask ventilation both without a leak (3.0 particles.l^-1^ (0 – 9 [0-43])) and with an intentional leak (11 particles.l^-1^ (7.0 – 26 [1-62])) was 64-fold and 17-fold lower than that of tidal breathing (p=0.001 and p=0.002 respectively). The peak particle concentration during facemask ventilation both without a leak (60 particles.l^-1^ (0 – 60 [0-120])) and with a leak (120 particles.l^-1^ (60 – 180 [60-480]) were respectively 20-fold and 10-fold lower than a cough (1260 particles (800 – 3242 [100-3682]), p=0.002 and p=0.001 respectively). This study demonstrates that facemask ventilation, even performed with an intentional leak, does not generate high levels of bioaerosol. On the basis of this evidence, facemask ventilation should not be considered an aerosol generating procedure.

## Introduction

The COVID-19 pandemic continues to place unprecedented demands on healthcare globally. There is evidence for airborne transmission of SARS-COV-2 [1–3], with natural respiratory activities and singing frequently implicated [4,5]. The World Health Organisation (WHO) and Public Health England (PHE) have focussed predominantly on droplet precautions to limit viral transmission for patients with a confirmed diagnosis of COVID-19. The use of airborne personal protective equipment (PPE) has been largely reserved for healthcare workers undertaking medical procedures deemed to be aerosol generating [6–8]. These procedures are presumed to generate as much or higher levels of bioaerosols from a patient’s respiratory tract than coughing and consequently carry an increased risk of viral transmission. The evidence for these putative ‘aerosol generating procedures’ is predominantly epidemiological and originates from the SARS epidemic in 2003 [9,10]. Several recent studies have questioned whether these medical procedures should be classified as ‘aerosol-generating’ following quantitation of the aerosol produced during these patient care activities [11–15].

Facemask ventilation is a core airway intervention currently on the WHO list of procedures deemed to be aerosol generating [9,10]. The epidemiological evidence for this designation is from a single study [16] that reported an increased risk of SARS infection from facemask ventilation before tracheal intubation. This risk was determined following interviews with 26 healthcare workers (an average 4 months after contracting SARS) to identify their clinical activity during the period 24 hours prior and 4 hours post-intubation. The authors reported a pooled increased risk of infection after being in the room of a SARS positive patient during intubation, where facemask ventilation had been performed (odds ratio 2.8 (95% CI 1.3-6.4)) [9,16]. Twenty three of these 26 healthcare workers were infected by just 3 patients and it should be noted the authors found performing an electrocardiogram was associated with an even higher risk of SARS infection (odds ratio 3.5 (95% CI 1.58-7.86)).

No study to date has set out specifically to measure the aerosol generated during facemask ventilation. Two recent clinical studies, performed in operating theatres, demonstrated a lack of aerosol generation for laryngoscopy and tracheal intubation [11,17]. Analysis during the phase of facemask ventilation of the anaesthetised patient, prior to tracheal intubation, demonstrated conflicting results. Brown *et al*. (2021) [11] reported that facemask ventilation was not aerosol generating. In contrast, Dhillon et al. (2021) [17] recorded an increased particle concentration above background during a period including facemask ventilation. Resolution of these different findings is of crucial importance as the ability to ventilate a patient via a facemask is a key component of both elective and emergency airway management. We therefore co-developed an experimental protocol to test specifically whether facemask ventilation is a high-risk procedure for aerosol generation. To assess the relative risk from facemask ventilation, we measured aerosol generation during facemask ventilation and compared this against each patient’s own tidal breathing and volitional coughs.

## Methods

Ethical approval was granted by the Greater Manchester REC committee (Reference: 20/NW/0393) as part of the AERATOR study (approved 18/09/2020). The study was granted Urgent Public Health status by NIHR and is registered in the ISRCTN registry (ISRCT:N21447815). The methods for aerosol measurement have previously been described in detail [11]. In brief a prospective environmental monitoring study was conducted in operating theatres in a UK hospital (Southmead Hospital, North Bristol NHS Trust).

All recordings were made within operating theatres with an ultraclean ventilation system (EXFLOW 32, Howorth Air Technology, Farnworth, UK) placed in standby mode as described in detail previously [18,19]. This provides an environment with very low background aerosol concentrations, an air change rate of 25 per hour (in line with most other operating theatres in the UK), an air velocity of 0.25m.s^-1^ at 1m above the ground, an air temperature of 20°C and humidity between 40-60%. An optical particle sizer (OPS) (TSI Incorporated, model 3330, Shoreview, NM, USA) was used to record particle size, concentration and mass (within the size range 300 nm to 10 µm in diameter) at a sampling rate of 1 Hz. A 3D-printed funnel was formed of polylactic acid on a RAISE3D Pro2 Printer, (3DGBIRE, UK) with 90 mm height, a 10 mm exit port and maximum diameter of 150 mm. This sampling funnel was connected to the OPS by a 1.25 m length of conductive silicone tubing of 4.8□mm internal diameter.

All consented participants were over 18 years of age, American Society of Anesthesiologists (ASA) physical status grade 1 or 2, undergoing routine elective surgery requiring neuromuscular blockade prior to tracheal intubation and had a negative COVID-19 polymerase chain reaction (PCR) test in the previous 72 hours. Patients with symptomatic gastro-oesophageal reflux, a potential or known ‘difficult’ airway, ASA 3 or greater or of body mass index (BMI) > 40kg.m^-2^ were excluded. The anaesthetic and theatre team undertook their normal practice during airway management with the single exception of intentionally generating a leak from the facemask during patient ventilation, after induction of anaesthesia (as detailed below). The researchers were not involved in the delivery of anaesthetic care. All staff in theatre wore airborne precaution PPE including FFP3-type masks. All participants were supine with head positioning as per the anaesthetist’s preference. An initial period of aerosol sampling was recorded, with the patient awake for 60 seconds of tidal breathing followed by three volitional coughs spaced at 30 second intervals. Sampling was performed with the funnel at a distance of 20 cm directly above the patient’s mouth. A piece of sampling tubing was cut to 20cm length and used to guide funnel positioning for each patient. The sampling funnel was then directed away from the patient to record background aerosol concentration in theatre whilst the anaesthetist prepared the patient for induction of anaesthesia.

All patients were pre-oxygenated with 100% oxygen, had intravenous induction of anaesthesia with propofol and an opioid followed by neuromuscular blockade with rocuronium (0.4-0.6 mg.kg^-1^). Once the patient was unconscious, facemask ventilation was performed via an anaesthetic circuit (circle system) connected to the anaesthetic machine (Aisys CS^2^, GE Healthcare, USA) enabling measurement of both the airway pressure and volume of manual breaths delivered. To ensure standardisation manual facemask ventilation was performed with a tidal volume of 5-7 ml.kg^-1^, airway pressures below 20 cm H_2_O and a respiratory rate of 12-15 breaths per minute. Standard anaesthetic monitoring, including waveform capnography and pulse oximetry, was used throughout. No airway adjuncts (e.g. Guedel airways) were required.

During the 60 seconds of ‘standard’ facemask ventilation, sampling was performed with the funnel at a distance of 20 cm directly above the patient’s mouth. If the patient was clinically stable and their lungs easy to ventilate, the anaesthetist then relaxed their grip on the mask to create an intentional, audible airway leak at the patient-mask interface. The patient’s lungs were then ventilated with the intentional leak for a further 60 seconds. The fresh gas flow and pressure limiting valve were adjusted to ensure the bag refilled to allow manual ventilation during this period. Monitoring of oxygen saturation was undertaken to ensure it did not fall below 95% (which would trigger restoration of ventilation with a good seal). During facemask ventilation with an intentional leak, the sampling funnel was moved towards the side of the facemask, directed towards the leak, maintaining a distance of 20 cm from the patient’s mouth. The funnel was handheld to ensure it could be promptly removed in case of clinical need.

Airway management events were timestamped by the researcher including: the period of tidal breathing, coughs, induction of anaesthesia, administration of neuromuscular blockade, start of recording for facemask ventilation with no leak and start of facemask ventilation with a leak. Facemask ventilation analysis was commenced ∼60 seconds after rocuronium had been administered. Aerosol sampling was continuous throughout the whole period from induction of anaesthesia to intubation.

Data were processed in the TSI Aerosol Instrument Manager software, and analysed in Origin Pro (Originlab, Northampton, Massachusetts, USA) and Prism v9 (Graphpad, San Diego, USA). The normality of the data distribution was assessed using the Shapiro-Wilk test. Comparisons were made between aerosol measurements with parametric or non-parametric statistical analyses (as appropriate and as indicted in the text). The significance level was set at P < 0.05. All data are presented as mean (SD) or median (IQR [Range]).

## Results

Recordings were made during airway management for 11 patients undergoing elective surgery. There were six women and five men with mean age 60 (18.7) and mean BMI 27.1 kg.m^-2^ (5.0).

The ultraclean ventilation system in the theatre environment produced a very low background particle concentration (median 2.1 particles.l^-1^ (0.7-4.6 [0-12.9]), n=11). Spontaneous quiet tidal breathing was consistently detected above background levels (Figure 1) with a median particle concentration of 191 particles.l^-1^ (77-486 [3.8-1313]) (n=11, p=0.002, Wilcoxon matched-pairs) (Figure 2A).

**Figure 1.**
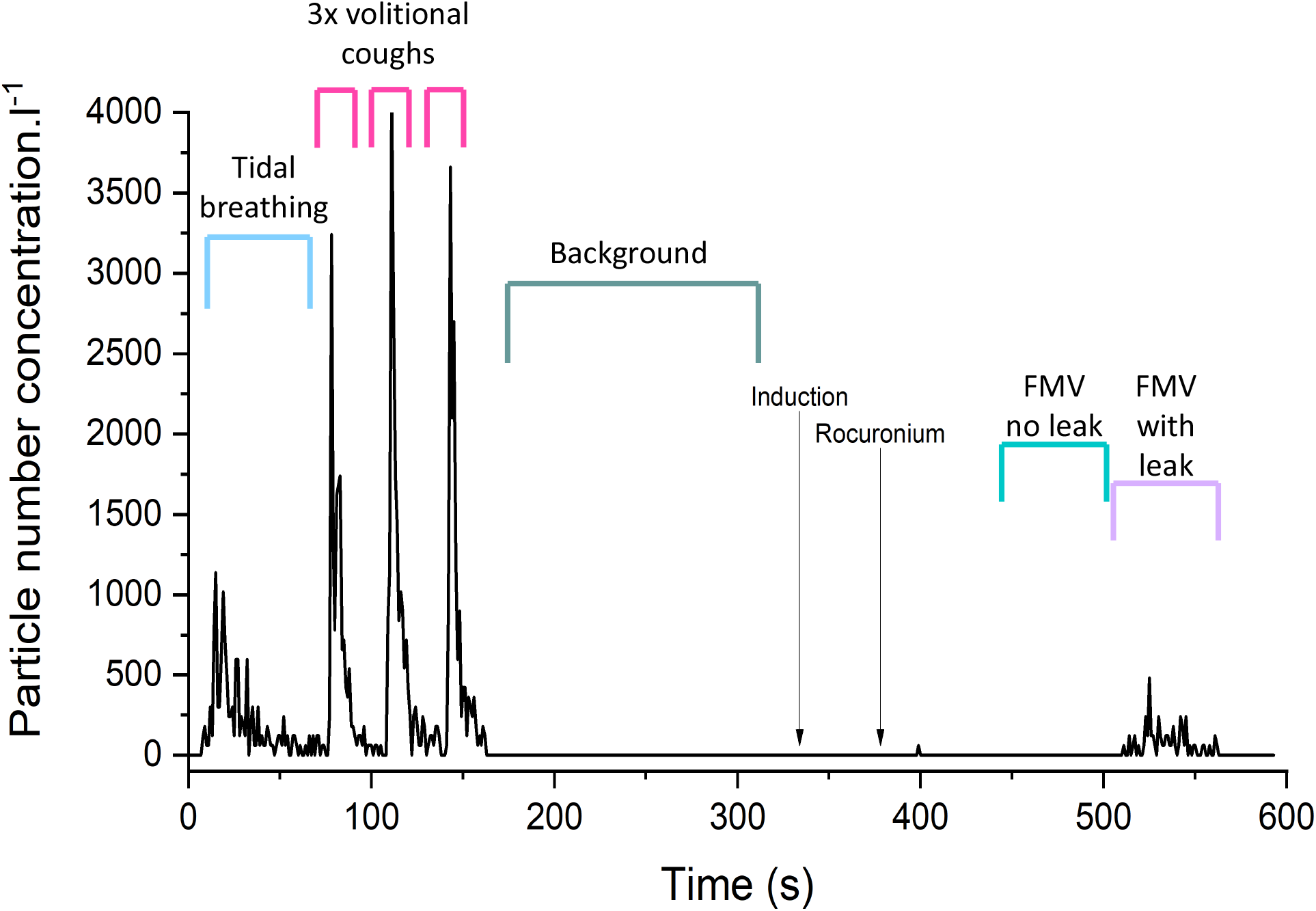
Aerosol concentration measured during the experimental protocol. This shows the number concentration of particles detected during baseline respiratory manoeuvres (tidal breathing and voluntary coughs), background monitoring, facemask ventilation (FMV) with no leak, and facemask ventilation with a leak.

**Figure 2.**
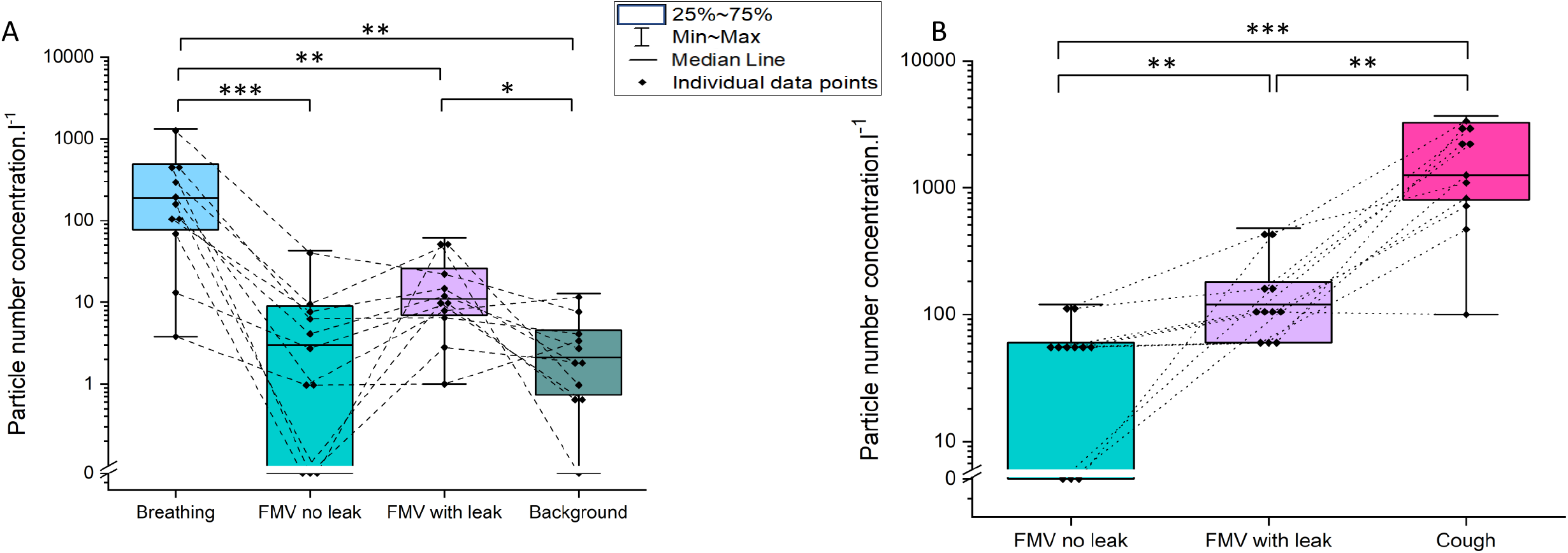
**A**. Comparison of particle number concentrations detected during tidal breathing, facemask ventilation (FMV) with/and without a leak and background levels. Dotted lines link values for each participant. **B**. Peak particle concentrations from facemask ventilation with/and without leak and cough. Wilcoxon matched pairs, *** p ≤0.001, ** p ≤0.01, * p ≤0.05)

The volitional coughs showed a median peak aerosol concentration of 1260 particles.l^-1^ (800-3242 [100-3682]) (n=11). Analysis of the particle size distributions of these coughs demonstrated the distinctive ‘fingerprint’ of a cough observed in other studies with the majority of particles (86.5%) less than 1 µm diameter[11,18-20]. These coughs also had a characteristic temporal profile with a rapid increase in particles which decayed over the subsequent 15 seconds.

No airway difficulties were experienced, and no request was made by the anaesthetists to remove the sampling funnel from the airway management zone. Oxygen saturation remained above 95% for all participants during facemask ventilation including during the period of ventilation with an intentional leak. There were no coughs noted during facemask ventilation either with or without a leak. The median particle concentration detected during the 60 s of facemask ventilation without a leak was 3.0 particles.l^-1^ (0 – 9 [0-43]) which was no different to background (2.1 particles.l^-1^, p=0.43) and much lower than the concentration recorded during tidal breathing (191 particles.l^-1^, p=0.001, Wilcoxon matched-pairs) (Figure 1 & 2A).

The median particle concentration during facemask ventilation with a leak was 11 particles.l^-1^ (7.0 – 26 [1-62]) which was around 5-fold higher than background (2.1 particles.l^-1^, p=0.019) but still much lower (17-fold) than that seen during tidal breathing (191 particles.l^-1^, p=0.002, Wilcoxon matched-pairs). Analysis of the difference in particle concentration between facemask ventilation with and without a leak showed no statistically significant difference (3 v 11 particles.l^-1^, p=0.074, Wilcoxon matched-pairs) (Figure 2A).

The peak particle concentration recorded during the periods of facemask ventilation without a leak was 60 particles.l^-1^ (0 – 60 [0-120]) and with a leak 120 particles.l^-1^ (60 – 180 [60-480]) and respectively 20-fold and 10-fold lower than the particle count detected during a cough (1260 particles (800 – 3242 [100-3682]) (p=0.002 and p=0.001 respectively, Wilcoxon matched-pairs) (Figure 2B).

## Discussion

This study demonstrates that facemask ventilation in anaesthetised patients, even with a leak, generates less aerosol than the patient’s own tidal breathing and far less aerosol than a cough. This supports the findings from our previous study which included periods of facemask ventilation as part of the intubation sequence [11]. We find no evidence that the procedure of facemask ventilation in these circumstances generates high concentrations of aerosol and therefore it should not be classified as an ‘aerosol generating procedure’ because it does not meet the definitions of such [6,21]. This has implications in a wide range of settings including during routine anaesthetic airway management. The avoidance of facemask ventilation before tracheal intubation or supraglottic airway insertion, due to concerns around aerosol generation, is not supported by our evidence and likely serves only to increase the risk of encountering difficulties in airway management.

We have used the patient’s own tidal breathing and coughs to enable within-subject comparison and relative risk estimation for facemask ventilation. Considerable inter-person variation in aerosol generation was identified in this study – with interpersonal variation ranging up to 50-fold for tidal breathing and 36-fold during coughing. This is in keeping with previous studies performed by the AERATOR group and others [11,12,15,18,19,22]. Therefore, using each participant as their own reference increases the power to generate meaningful comparisons from a relatively small sample. We have also modified our aerosol sampling position to move from 0.5 m to 0.2 m to be closer to the patient’s mouth. This has increased our ability to detect the emitted aerosol from source and has increased the concentration of particles recorded with tidal breathing and other respiratory activities. There was a 48-fold increase in particle concentration detected during tidal breathing when recorded at this closer position (191 particles.l^-1^ vs 4 particles.l^-1^ - from our previous study of supraglottic airways in the same environment [19]). The higher measured particle concentrations closer to the mouth are likely due to decreased particle dispersion and the capture of particles with low momentum in the smaller size range when sampling at 0.2 m compared to 0.5 m. Despite sampling in closer proximity to the source it is possible some aerosol was not detected during facemask ventilation but as the concentration detected was far lower than that produced by each patient’s own breathing, we can infer the relative risk of aerosol generation by facemask ventilation is very low.

The low concentration of aerosol detected during facemask ventilation with an intentional leak is also reassuring given that this probably represents a worst-case scenario. The particles detected during face mask ventilation with leak likely represent respiratory aerosol originating from the lungs and upper airways during continued positive pressure ventilation rather than from turbulent airflow over the face. This is supported by the fact that these particles were predominantly small – which is consistent with respiratory origin where the smallest particles are thought to be generated [23]). We emphasise however that this concentration of aerosol was far lower than the patient would generate if conscious and breathing at rest. We can extend this conclusion further to state that a well-fitting facemask with a good seal reduces emitted aerosol concentration to the point where it is indistinguishable from the near-zero aerosol background, and we have previously demonstrated a well-fitting facemask can prevent bioaerosol leak from a cough [19] by keeping respiratory aerosols within the anaesthetic circuit. This is entirely predictable as the mask forms a physical barrier to aerosol spread.

A limitation of the study is that we intentionally studied a period of facemask ventilation after neuromuscular blockade in order to focus on the aerosol generation associated with the specific procedure rather than any paroxysmal respiratory event like coughing (as per the PHE interpretation of the AGP definition). However, aerosol sampling was conducted continuously throughout induction of anaesthesia and the period of facemask ventilation performed immediately before the formally analysed period (ie before neuromuscular blockade) also did not show increased aerosol concentrations above background (Figure 1). Previous work performed by our group measured aerosol generation during facemask ventilation in anaesthetised patients without neuromuscular blockade, which again did not demonstrate aerosol generation [19]. So we are confident that our conclusions may be generalisable to the unparalysed patient.

Very low background particle counts, accurate timestamping and a high detector sampling rate are essential for accurate detection and attribution of aerosol from respiratory events and medical procedures. During the conduct of this study, we did not detect any non-attributable spikes of aerosol. We have previously identified a variety of materials present in an operating theatre capable of generating high levels of (non-respiratory) airborne particles including patient bedding, gauze swabs, tube-ties, throat packs, surgical scrubs and incontinence pads [19]. The study by Dhillon and colleagues, which reported episodes of increased aerosol during the period of facemask ventilation [17] was not performed in a theatre with an ultraclean ventilation system making precise source attribution more challenging. There may also have been other procedures conducted during the induction sequence that could have generated aerosol, such as airway suction (currently considered an AGP which also requires further exploration).

This study demonstrates facemask ventilation, even performed with an intentional leak, does not generate high levels of bioaerosol. Both tidal breathing and a volitional cough generate many-fold more aerosol than facemask ventilation. On this basis we believe facemask ventilation should not be considered an aerosol generating procedure. These findings, along with previous studies reporting that tracheal intubation/extubation and supraglottic airway insertion/removal are not aerosol generating procedures, should reassure practitioners about the relative risks associated with these airway management strategies.

Accumulating evidence demonstrates many procedures currently defined as AGPs are not intrinsically high risk for generating aerosol, and that natural patient respiratory events (such as coughing) often generate far higher levels [11–14]. Furthermore, some of those procedures that do generate aerosol (such as oesophagogastroduodenal endoscopy) only do so when they cause coughing [18]. The emerging evidence from quantitative clinical aerosol studies is yet to be incorporated into clinical guidance for aerosol generating procedures and we believe this needs urgent re-assessment. Declassification of some of these anaesthesia-related procedures as ‘Aerosol Generating Procedures’ would seem appropriate due to their lack of aerosol generation. Our findings also raise the broader question of whether the term AGP is still a useful concept for anaesthetic airway management practice in the prevention of SARS-COV-2 or other airborne pathogens [24].

## Data Availability

Data available on request from the authors

## Acknowledgments

The AERATOR study is registered in the ISRCTN registry (ISRCT:N21447815). Andrew Shrimpton is an NIHR funded doctoral Research Fellow, NIHR301520. Bryan Bzdek is supported by the Natural Environment Research Council (NE/P018459/1). Fergus Hamilton is a Wellcome GW4 funded Clinical Doctoral Fellow. AERATOR is funded by an NIHR-UKRI rapid rolling grant (Ref: COV0333). *This report presents independent research commissioned by the National Institute for Health Research (NIHR). The views and opinions expressed by authors in this publication are those of the authors and do not necessarily reflect those of the NHS, the NIHR, UKRI, or the Department of Health*.

